# Facial photographs reveal mortality risk beyond triage

**DOI:** 10.64898/2026.02.12.26346148

**Authors:** Indra Heckenbach, Kasper Karmark Iversen, Michael Ben Ezra, Mikkel Porsborg Andersen, Henning Bundgaard, Morten Scheibye-Knudsen, Rasmus Bo Hasselbalch

## Abstract

Rapid risk stratification is essential in the clinic, yet vital signs, laboratory tests, and triage scores may not fully capture risk at presentation. We investigated whether facial photographs taken after emergency admission provide an additional mortality signal. Using 27,660 smartphone facial photographs, we trained deep neural networks to identify mortality risk with a Cox proportional hazards framework. Face-derived risk scores strongly stratified short- and long-term mortality, outperforming triage levels, vital signs, blood chemistry, and facial age estimation. Face scores revealed substantial heterogeneity in risk within each triage category, including low-acuity patients with mortality risk exceeding that of high-acuity levels. Combining high-acuity triage with high face score quartiles identified patients with a sixty-fold increased mortality risk within 30 days (odds ratio 60.43 vs lowest risk and triage), while triage alone resulted in six-fold increase in mortality (odds ratio 6.05 vs lowest triage). Integrating facial risk scores with routine clinical variables further improved risk stratification. These findings indicate that unstructured facial photographs encode clinically actionable risk and may augment existing triage and monitoring strategies.

## Introduction

Medicine relies on rapid risk stratification to optimize patient outcomes in many settings^1–4^. Traditional tools such as the patient’s vitals and blood chemistry panels are essential for assessing risk and guiding triage and treatment^5,6^. However, clinicians may also infer patient health from subtle cues, often referred to as the gestalt, which can help inform observation, treatment and follow-up^7,8^. Optimizing clinical evaluation has significant potential to improve patient outcome^9–11^. Deep learning can provide objective, quantified assessment of clinical signs, augmenting established methods.

The human face encodes age, emotional, and physiological conditions^12,13^, and could provide a rich source of clinical information. Recent work in machine learning has led to “aging clocks” that estimate chronological age from sources like DNA methylation and proteomics but also from human facial photographs^14–16^. Notably, age residuals (deviations from chronological age) are associated with obesity, blood pressure, and cholesterol levels and have emerged as a potential biomarker of heart disease, kidney disease, diabetes, stroke and hypertension^17,18^. Yet, the prognostic utility of facial features in clinical settings such as emergency departments is unexplored. Most prior facial-image studies infer mortality through age-acceleration models rather than training on survival signals directly, and none have assessed prognostic value in acute care.

In this study, we investigated whether facial photos taken after emergency admissions can be used to identify risk. Our dataset constitutes the first large, real-world clinical cohort enabling such evaluation. Here we show that a single non-standardized smartphone photograph of the face, taken without any protocol, identifies mortality beyond age, vital signs, blood tests, or triage level. This approach introduces a new modality for rapid, non-invasive risk assessment in the clinic.

## Results

### Face-derived risk stratifies short- and long-term mortality

Using facial photographs from 27,660 unique patients hospitalized in Denmark (with 14,127 deaths over eight years), we developed deep learning models to identify mortality risk (Table 1, Extended Data Fig 1a). Face-derived risk scores were higher among patients who died sooner and differed by sex, and across a broad age range higher scores consistently tracked shorter survival, indicating detection of prognostic features beyond chronological age (Extended Data Fig 1b,c). Building on this signal, face scores provided a much stronger indicator of mortality risk and superior separation over time compared to the five-level Danish DEPT triage system^10^ (Fig 1a), with median survival decreasing from 1810 days in the lowest quartile to 341 days in the highest, a 5.3-fold reduction (Fig 1a). Face scores stratified mortality risk beyond triage levels, including within critical, urgent, non-urgent categories, revealing substantial heterogeneity in mortality risk among patients assigned the same triage level, with patients classified as low-acuity exhibiting face score risk often exceeding those of higher-acuity groups (Fig 1b). Face scores were highly effective at identifying high risk patients at all triage acuity levels, including those evaluated as low acuity. Receiver operating characteristic (ROC) analysis demonstrated strong discrimination for early mortality (AUC= 0.81 for ≤3-day mortality; Fig 1c). Patients in the top decile of age-adjusted face scores had an odds ratio (OR) of 5.39 [95% CI: 4.19-6.92, P<0.0001] for 3-days mortality compared with the lower 90% (Extended Fig 1d).

**Table 1:**
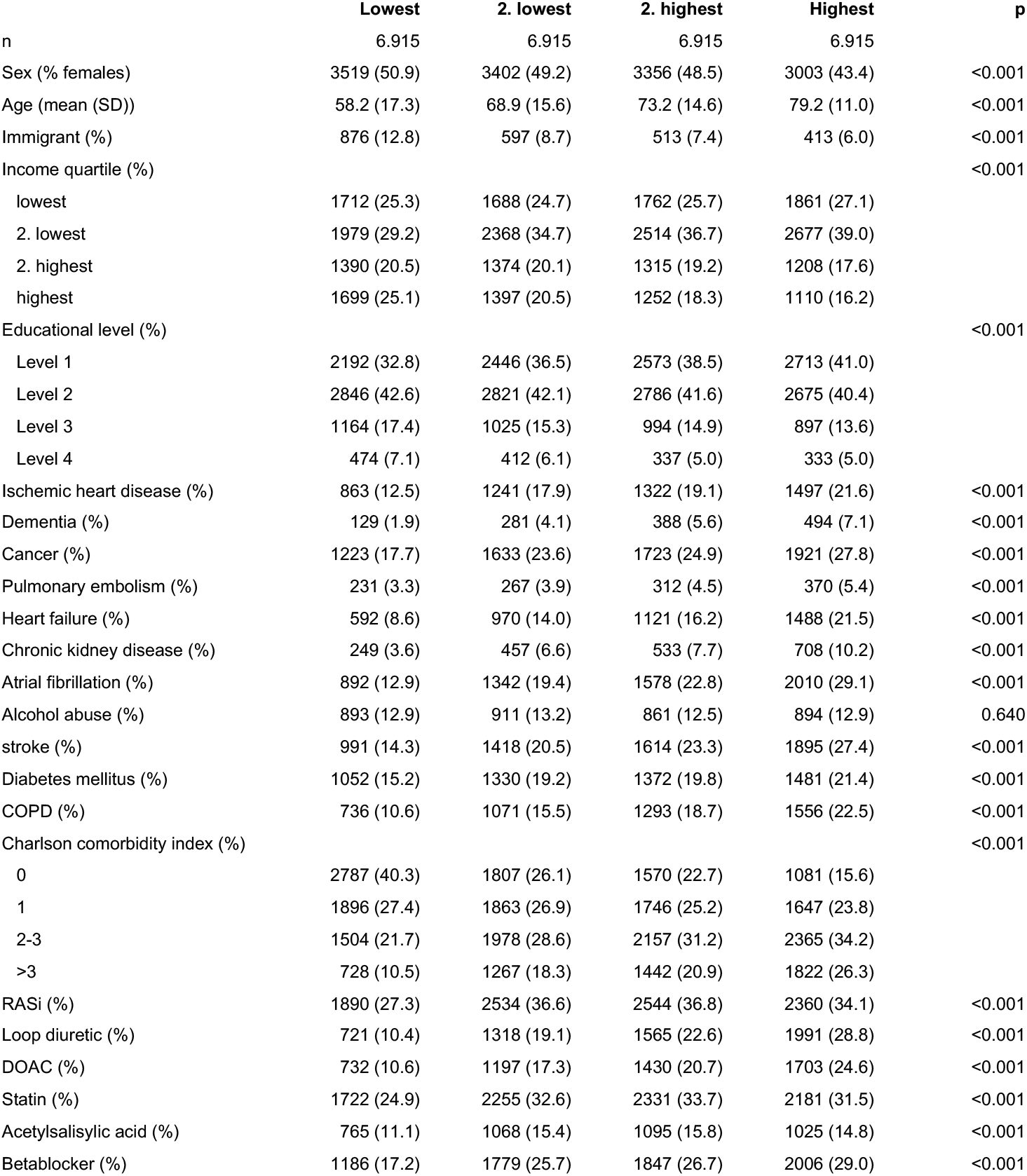
Patient characteristics. Characteristics of study participants, stratified by quartiles of predicted risk from multi-modal model (face score, biochemistry, and patient vitals). COPD: Chronic Obstructive Pulmonary Disease, DOAC: Direct Oral Anticoagulant, RASi: Renin-Angiotensin II inhibitor.

**Figure 1:**
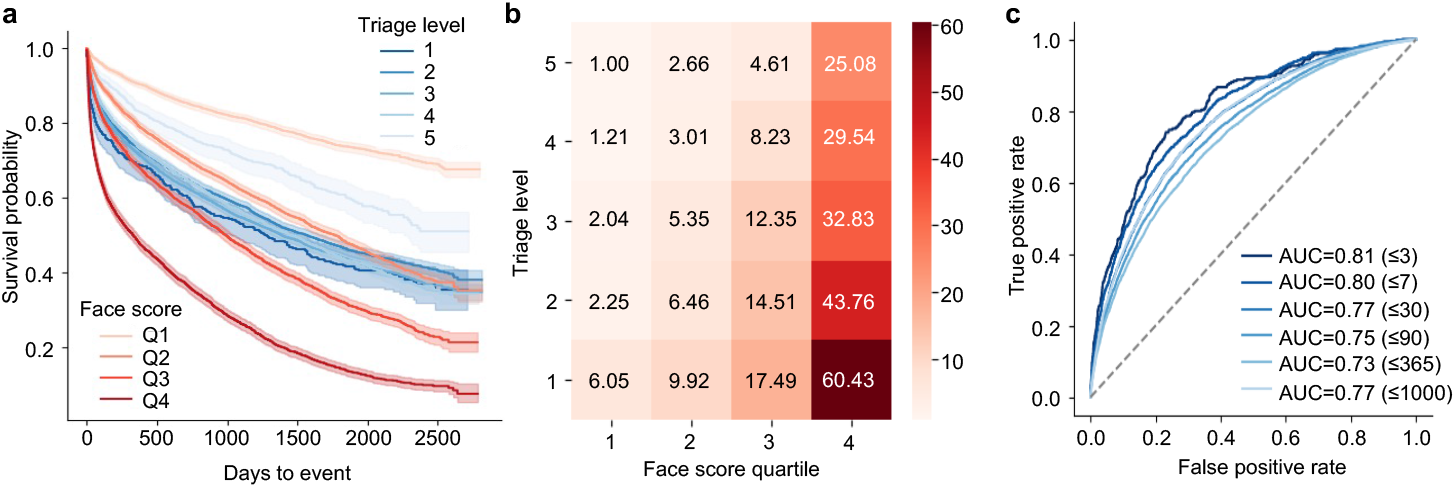
Facial photos reveal mortality risk beyond triage. **a**, Kaplan-Meier survival curves for quartiles of face scores and triage levels. **b**, Odds ratios of 30-day mortality by face score quartiles and triage levels, relative to reference (Q1/level 5). **c**, ROC performance of face score by survival time (in days).

### Mortality signals beyond accelerated age

To assess whether face scores reflected accelerated aging rather than independent prognostic information, we trained neural networks to estimate chronological age from facial photographs (Extended Data Fig 1e,f). Deviations between predicted and chronological age were associated with mortality risk, with prematurely aged appearances corresponding to increased risk and shorter survival (Extended Data Fig 1g,h). However, after adjusting face scores for chronological age or predicted age, substantial stratification of survival persisted, with the highest-risk quartile exhibiting markedly reduced survival compared with the lowest (Extended Data Fig. 1i, j). Stratified by age groups to reduce age effects, ROC analysis showed AUCs ranging from 0.71 to 0.79 for 7-day survival (Extended Data Fig 1k). Age-adjusted face scores showed substantial risk stratification across triage levels (Extended Data Fig 1l). These findings indicate that face-derived risk captures prognostic signals beyond facial age estimation and is not explained by accelerated aging.

### Comparison to triage

To determine the effectiveness of face scores in the clinic, we compared them to the 5-level Danish triage system (DEPT)^10^. Overall, face scores were significantly higher for critical triage level 1 compared to urgent to moderate levels 2-4, which had overlapping face scores, and face scores were significantly lower for non-urgent triage level 5 (Extended Data Fig 2a). For each triage level, we stratified those patients by age-adjusted face score quartiles (from the entire cohort), revealing substantial separation in mortality risk for each triage level (Extended Data Figs 2b-f). We also evaluated mortality rate per age-adjusted face score quartile by triage level for several time periods, showing a substantially higher mortality rate for the highest face score quartile versus lower quartiles for all triage levels and time points (Extended Data Fig 2g). Remarkably, the mortality rate for the highest face score quartile for non-urgent level 5 triage exceeded the lower three face score quartiles for other triage level patients, across all time points evaluated, meaning patients with the highest faces scores at the non-urgent triage level had higher mortality risk than patients at urgent and critical triage levels. These results demonstrate the face score provides insight into short- and long-term mortality risk well beyond established triage levels.

### Mortality risk from blood chemistry and vital signs

Patient blood test results and vitals were used to assess mortality risk. A Cox model, trained with blood chemistry alone, showed the highest contributing factors were concentrations of albumin, hemoglobin, potassium, CRP, neutrophils, and creatinine by quantifying each factor’s relative importance through removal of the factor and measuring the effect on the prediction (Extended Data Fig 3a). To investigate the impact of individual factors, separate Cox models were fitted to each factor plus age, and Kaplan-Meier curves were generated to illustrate their risk by quartiles (Extended Data Figs 3b-i). In addition, a Cox model based on patient vital parameters identified as the highest mortality contribution from weight, diastolic blood pressure, respiration rate, and saturation (Extended Data Fig 4a). Cox models were prepared for each factor along with age and used to evaluate survival curves by quartile (Extended Data Figs 4b-i). Combining blood factors, vitals, age, face score, and predicted age residuals, a new Cox model showed chronological age was the strongest risk factor, followed by face score, and then other key factors including age residual, weight, height, blood pressure, albumin, hemoglobin, neutrophils, and creatinine (Fig 2a). To evaluate their associations with patient age, a new age clock was trained on common blood chemistry factors, yielding an MAE of 9.71 years (RMSE=12.75 years, Extended Data Fig 4j). After vitals such as height, weight, blood pressure, temperature, etc. were included, an extended blood and vitals clock gave an MAE of 8.26 years (RMSE=10.92 years, Extended Data Fig 4k).

### Multi-modal survival models improve risk stratification

Face scores, predicted and chronological ages, biochemistry, and vital parameters were all combined to build an integrated survival model, which demonstrated higher risk for patients with shorter survival time for patients over 30 years (Extended Data Fig 5a). Kaplan-Meier survival analysis showed the highest-risk quartile has median survival of 104 days, compared to 689 days for the lower risk quartile (Extended Data Fig 5b). After adjusting for age (using residuals from a regression with risk score), Kaplan-Meier analysis showed the median survival of 892 days for the highest quartile compared to not reached for the lowest (Extended Data Fig 5c). ROC performance reached an AUC of 0.84 for ≤3-day survival, with a slight decline for longer timepoints (Fig 2b). Stratified by age groups for 7-day survival, ROC analysis showed AUC around 0.80 above age 40 (Extended Data Fig 5d). The top decile of scores versus the lower 90% had OR=8.81 [95% CI: 6.83-11.35, P<0.0001] for 3-day mortality (Extended Data Fig 5e).

**Figure 2:**
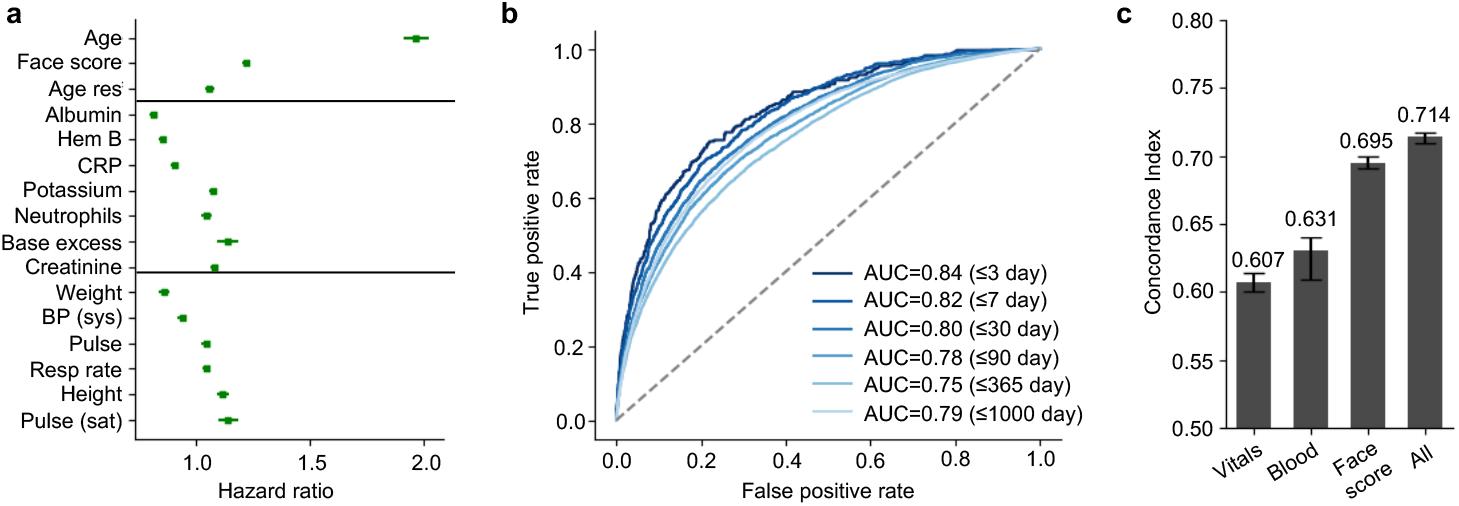
Multi-modal models improve mortality risk prediction. **a**, Risk contribution from blood, vitals, age, predicted age residual, and face score in a Cox survival model. **b**, ROC performance of face score, blood, vitals, age, and predicted aged by survival time. **c**, Concordance index by risk predictor model.

To compare how well the models identify risk, we calculated the concordance index (C-index) as a measure of the accuracy of the different models. Surprisingly, we found that face score alone had a C-index of 0.695 [95% CI: 0.691-0.700], suggesting that the model has considerable capabilities in risk stratifying patients and surpassing the models for blood factors and vital parameters with a C-index of 0.631 [95% CI: 0.609-0.640] and 0.607 [95% CI: 0.600-0.614], respectively. When face score, blood factors, vital parameters, age, and predicted age were all combined, the multimodal model gave slightly higher performance than face scores alone with a C-index of 0.714 [95% CI: 0.709-0.717].

### Facial features influence risk estimates

To better understand the visual signals that indicate survival, the face model was tested with colour-shifted images. Adjusting red and blue channels revealed reduced risk with more red and less blue, possibly indicating an effect of oxygenation on tissue hue (Extended Data Fig 5f). For red and green combinations, red decreased risk, while green increased risk (Extended Data Fig 5g). For green and blue, both together increased risk, possibly mimicking cyanosis through deoxygenated pallor (Extended Data Fig 5h). Additionally, facial images were evaluated with regions obfuscated and risk assessed with AUC metrics. Masking of the mouth and eyes led to a modest reduction in AUC and nose and eye region led to a more substantial decline (Extended Data Fig 5i). These findings suggests that it is a combination of many factors that drive predictions.

## Discussion

In this study, we show that facial photographs captured during routine emergency care encode clinically relevant mortality risk that is not fully captured by standard triage, vital signs, or laboratory tests. While face-derived risk scores broadly stratified survival across the cohort, their most clinically relevant contribution was the identification of substantial heterogeneity in risk within established triage categories, including patients classified as low acuity who exhibited mortality rates comparable to higher-acuity groups. These findings indicate that facial risk scores capture latent physiological signals that would complement existing triage systems.

Facial photographs alone stratified mortality risk across multiple time horizons, with models trained directly on time-to-death substantially outperforming approaches based on chronological age or age acceleration. Although deviations between predicted and chronological age were associated with survival, age-based models provided limited risk stratification. Comparisons to triage levels further indicate that face-derived risk captures physiological signals not reflected in standard clinical assessments. Notably, these signals were extracted from non-standardized smartphone photographs captured without a protocol, supporting the feasibility of rapid, low-cost risk assessment that augments existing triage and monitoring strategies. This study was based on photographs captured by smartphone cameras in an unstructured manner without following a protocol or SOP, enabling rapid deployment in both clinics and resource-limited and underserved settings worldwide.

Indeed, our approach provides a direct method for mortality prediction from non-standardized facial photographs captured after emergency department admissions, providing prognostic value beyond blood tests and vitals. In contrast, other facial aging predictors, such as FaceAge^19^, utilize age acceleration to infer mortality risk in oncology. A recent study on 442,110 facial photos from Wikipedia to predict age, found that age acceleration predicts all-cause mortality for individuals, concluding that face age serves as a low-cost proxy for biological age in personalized medicine^20^. Additionally, the FAHR-FaceSurvival model^21^, fine-tuned on cancer radiotherapy photos, predicts survival of oncology patients. Unlike these and other methods focused on accelerated aging, the face scores presented here capture physiological cues in single photos from patients following acute admission, providing non-invasive, generalizable AI for emergency risk assessment.

Our face score approach has the potential for clinical application through smartphone-based tools that capture patient photos for rapid assessment of mortality risk. Similar image-based models have been developed for other disease specific areas such as in the evaluation of skin cancers^22^. The use of AI in emergency medicine has attracted growing interest in recent years, with several retrospective studies showing the potential for superior risk prediction compared to traditional triage systems^23^. Facial imaging represents a promising, easily implemented modality that could enhance early triage, and then be integrated with vital signs^24^, laboratory values^25^, and other diagnostic tests for comprehensive prognostic assessment. Beyond emergency care, this technology could be adapted for primary care, telemedicine or in rural areas, where acute blood samples are unavailable, or risk-stratified scheduling of follow-up visits, personalized treatment intensity, and optimizing resource allocation. Additionally, serial risk assessments could serve as an objective biomarker for monitoring disease progression and treatment response.

Several limitations should be considered. First, data was collected from Eastern Denmark including the Zealand regions which may limit generalizability, as outcomes could reflect region-specific care. Second, a main concern with deep learning architecture is that it is not readily explainable, and the visual features studied may not be broadly applicable without training on additional demographics across multiple hospital systems. Third, model performance has not been evaluated in non-emergency clinical and nonclinical environments. Despite these constraints, initial results suggest a potential for substantial clinical value.

In conclusion, we demonstrate that deep learning models can accurately identify mortality risk from facial photos. This represents a promising approach for rapid clinical risk assessment.

## Contributions

IH wrote the manuscript, created deep learning models, and analyzed the data. RBH edited the manuscript, facilitated data sharing, and analyzed patient data. MBE advised on statistical methods. MA advised the project and revised the manuscript. HB advised the project and revised the manuscript. KI revised the manuscript. MSK revised the manuscript, advised on deep learning and analysis, and supervised the project.

## Data Availability

Data supporting the findings of this study are available from the corresponding author upon reasonable request. Due to GDPR restrictions, individual-level facial images and linked clinical data cannot be shared. The study protocol, statistical analysis plan, and informed consent materials are available from the corresponding author on reasonable request.

## Code Availability

Code for model training and analysis will be made publicly available on GitHub upon publication.

## Acknowledgements

MS-K was supported by the Novo Nordisk Foundation (NNF0089176 and NNF17OC0027812) and the Danish Cancer Society (R368-A21521, R302-A17379_001, and R167-A11015_001).

## Reporting Summary

Further information on research design is available in the Nature Portfolio Reporting Summary linked to this article.

## Supplementary Information

Supplementary Information is available for this paper.

## Methods

### Data Source

In this study, we obtained individuals’ age, sex, and vital status from electronic medical records. Following admission to the emergency department, front-facing facial photographs were captured using smartphone cameras in a non-standardized manner, without adherence to a specific protocol or standard operating procedure (SOP) as a part of routine care. These images were stored in the patients’ electronic medical records (Epic). We included individuals with an image stored within 30 days of admission to an emergency department.

All Danish residents are assigned a unique civil personal registration number at birth or upon immigration, enabling linkage and collection of data from the Danish nationwide registers.

The Danish registries record information administratively on a national level for economic, social, and healthcare purposes. To determine comorbidities related to the image data we retrieved data on hospitalizations and related diagnoses since 1973 from the Danish National Patient Register^26^. Furthermore, we used The Danish National Prescription Register to obtain information on all redeemed prescription medications^27^.

### Patient Cohort

Facial photographs were obtained from 27,660 unique patients (52% male; median age, 73 years [range, 18-104]; Table 1) who had been admitted to emergency departments in Denmark between May 24, 2016, and May 14, 2024; photographs were obtained within 30 days of admission, and 14,127 patients died within the study period.

### DEPT Triage

The Danish emergency process triage (DEPT) is a five-level system used in Danish emergency departments to prioritise patients based on vital signs (oxygen saturation, heart rate, blood pressure, Glasgow Coma Scale, temperature, respiratory rate) and emergency concerns^10^. Level 1 is critical, requiring immediate physician contact, level 2 is very urgent, level 3 is urgent, level 4 is less urgent, and level 5 is non-urgent.

### Image Preparation

Smartphone camera photos were used to build a data set by isolating facial regions. In a random sample of 96 images the facial region was annotated, and a U-Net model was then trained to detect facial regions from the images. After segmentation of all images, the detected facial region of the image was extracted and resized proportionally to fit 299×299 pixel images. The full set of images was shuffled and split into 3 equal-sized sets for processing.

### Deep Learning Models

Several deep learning models were developed to predict chronological age and mortality risk, using 3-fold cross-validation training. Facial aging models were trained using Xception, mapping the segmented facial images to chronological age output using regression. Mortality risk was trained using EfficientB3, mapping facial images to predicted log hazard ratios using a Cox proportional hazards loss function, which leverages observed time-to-event (death), based on the DeepSurv approach^28^. The proportional hazards (PH) assumption of the Cox-based DeepSurv model was evaluated by visually comparing Kaplan-Meier curves across quartiles of predicted risk, with the absence of curve crossing indicating that the assumption was satisfied. Integrated models that utilized facial photos, age, vitals, and blood factors were developed with a multi-layer deep neural network with dropout that takes input factors as scaled numerical values and generates survival log hazard ratios using a similar Cox proportional hazards loss function. To further improve performance, an ensemble was used: age and risk models were trained 10 times with random initialization, and on inference each model was run separately and their predictions combined with a mean function to produce face scores.

### Statistics

Survival data was analyzed using Kaplan-Meier survival curves and Cox proportional hazards models to determine hazard ratios (HR). We tested associations between predicted risk and mortality events at several timepoints (3 days, 7 days, 30 days, 90 days, 365 days, and 1,000 days) using receiver operating characteristics (ROC) and compared the results using area under the curve (AUC). When blood chemistry and vitals were missing in some patients, Cox regression with multiple factors was done using indicator variables and imputation with mean values, and for single factor analysis patients with missing data were excluded. A p-value of <0.05 was considered significant. All analysis was done using Python 3.12.7, Statsmodels 0.14.4, Linelines 0.30.0, Scikit-learn 1.5.2, Scikit-survival 0.23.0, SciPy 1.11.4, Numpy 1.26.4. PyTorch 2.4.1, Torchvision 0.19.1, Timm 1.0.9.

### Ethics

The study has been presented to the National Ethics Committee who waived the need for ethical approval. The study is approved from the Record Data Team, Center for Health in the Capital Region of Denmark as well as been approved by the data responsible institute Capital Region of Denmark in line with the Danish Data Protection Act and the General Data Protection Regulation. Data from Danish registries were accessed through the servers of Statistics Denmark.

