## Extended Figures for "Facial photographs reveal mortality risk beyond triage"

### Extended Data Figures

#### *Contents*

**Extended Figure 1: Patient photos reveal mortality risk beyond the signs of aging** **2**

**Extended Figure 2: Face scores improve risk stratification beyond triage** **4**

**Extended Figure 3: Blood factors contribute to mortality risk** **5**

**Extended Figure 4: Vitals contribute to mortality risk** **6**

**Extended Figure 5: Face regions and color contribute to mortality prediction** **7**

### Extended Figure 1: Patient photos reveal mortality risk beyond the signs of aging

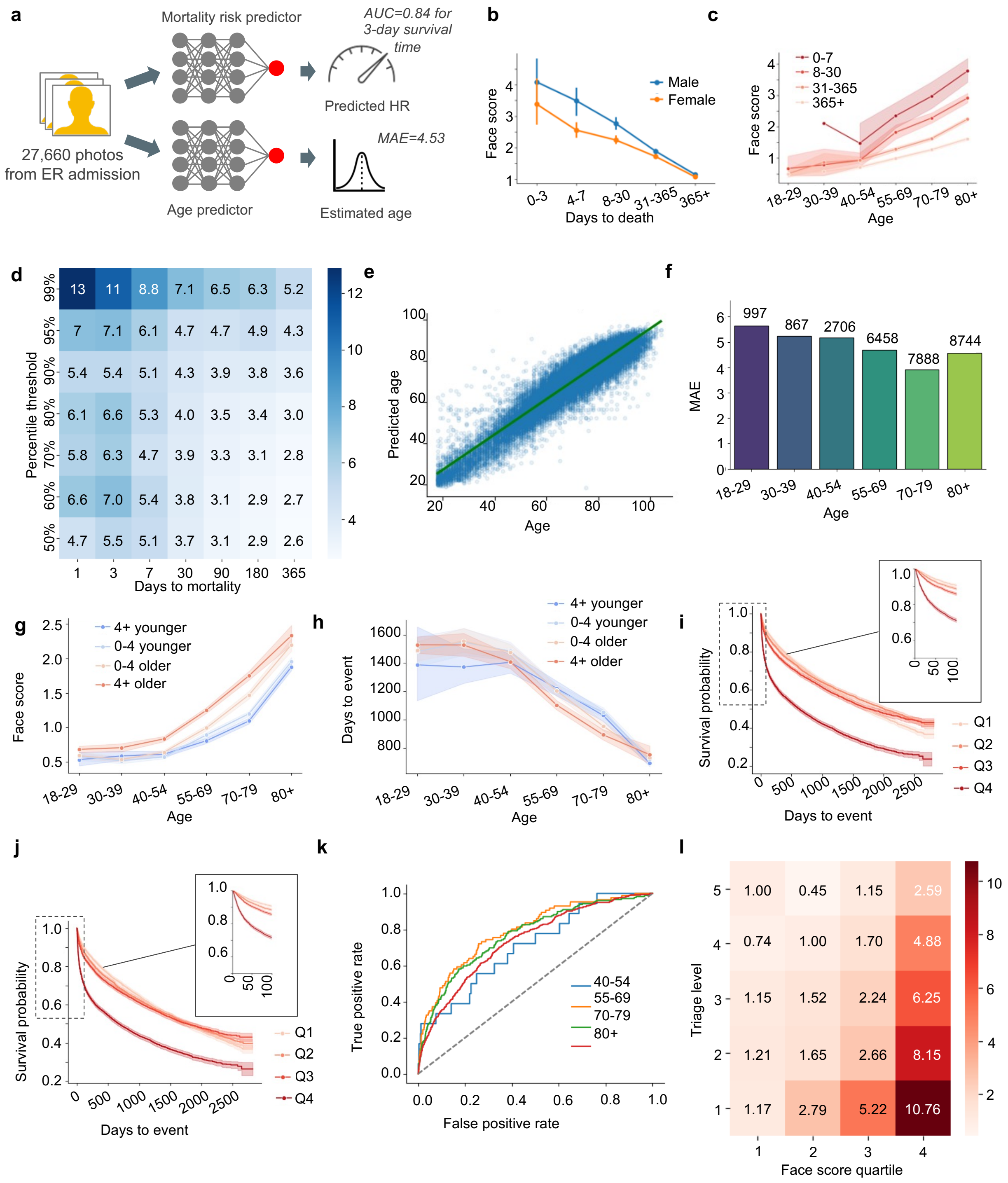


a, Workflow from facial photos to risk and age predictors. b, Mean HR scores by survival days. c, Mean HR scores by age range, stratified by survival time. d, Odds ratios based on face scores of upper percentiles vs lower for several time points. e, Age predicted from facial photos (MAE=4.53, RMSE=5.95, n=27660). f, Residuals of predicted age. g, Mean HR scores by age range, stratified by age residual groups. h, Survival time by age range, stratified by age residual groups. i, Kaplan-Meier survival curves by quartile of age-adjusted risk scores with expanded view. j, Kaplan-Meier survival curves by quartile of predicted age-adjusted risk scores with expanded view. k, ROC performance by age group for 7-day survival time (40-54: AUC=0.71, 55-69: AUC=0.79, 70-79: AUC=0.77, 80+: AUC=0.74). l, Odds ratios of 30-day mortality by age-adjusted face score quartiles and triage levels, relative to reference (Q1 face score and low-acuity triage level 5).

Extended Figure 2: Face scores improve risk stratification beyond triage

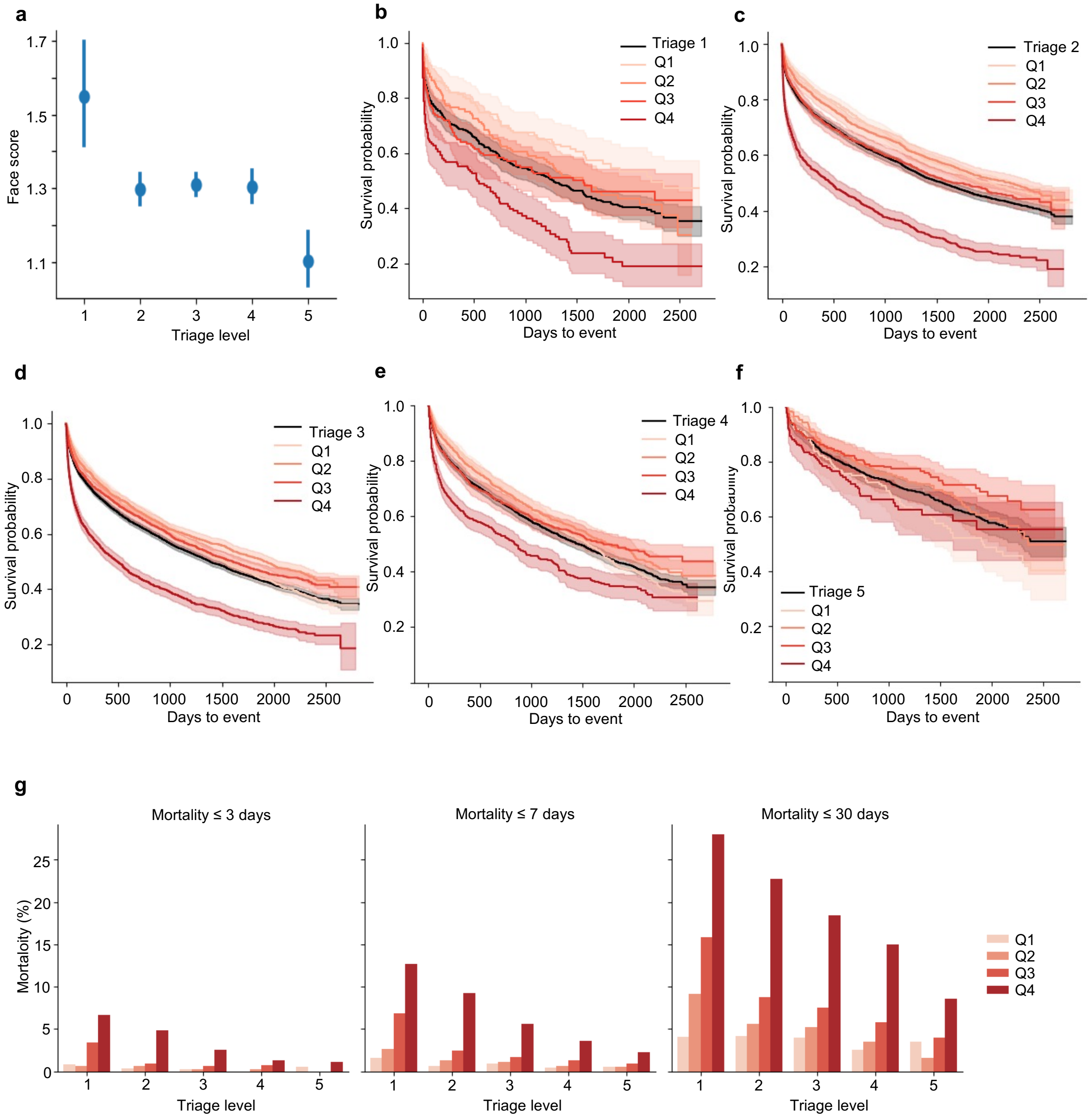


**a**, face score by triage level. **b**, Kaplan-Meier survival for triage 1 (black) and triage 1 stratified by risk quartile using age-adjusted face scores (reds). **c**, Kaplan-Meier survival for triage 2 (see b). **d**, Kaplan-Meier survival for triage 3 (see b). **e**, Kaplan-Meier survival for triage 4 (see b). **f**, Kaplan-Meier survival for triage 5 (see b). **g**, Mortality rate by age-adjusted face score quartile, stratified by triage level, for several tine points.

#### Extended Figure 3: Blood factors contribute to mortality risk

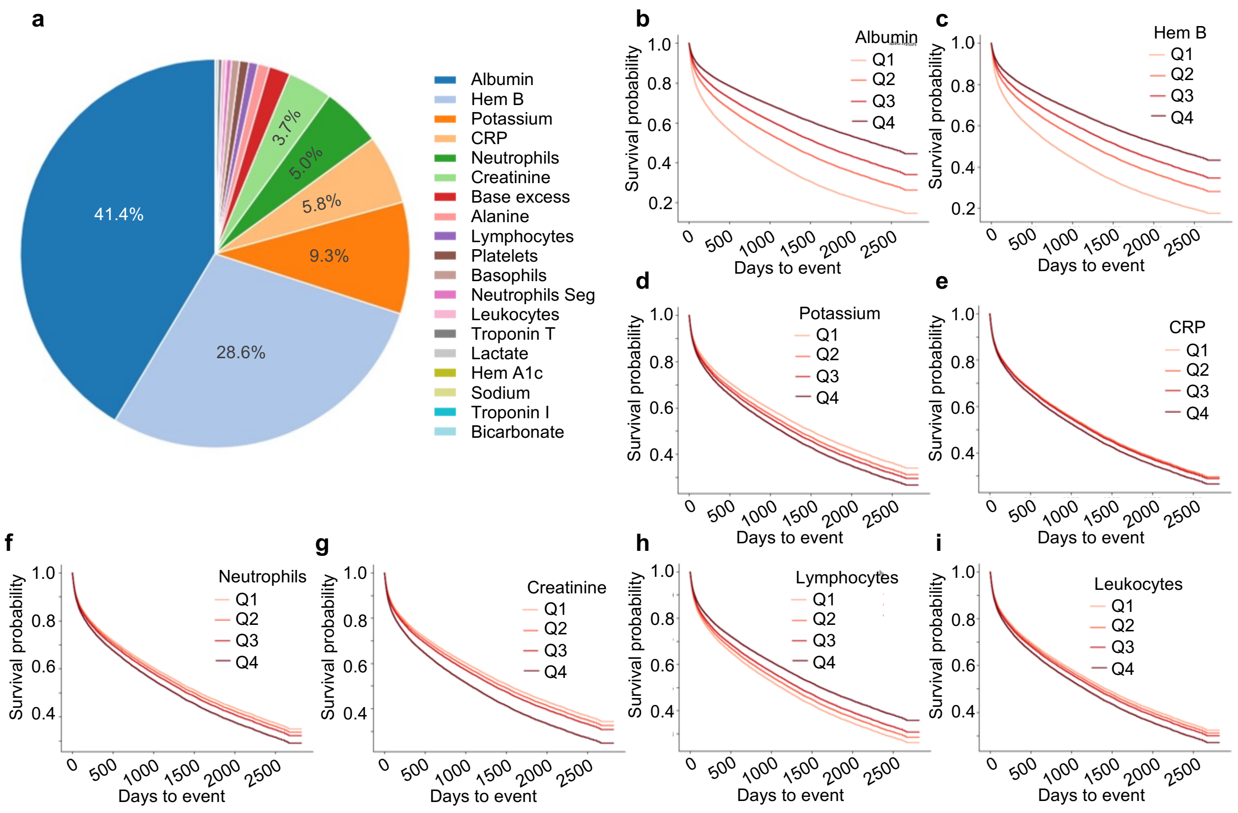


**a**, Breakdown of blood factors contributing to risk in a Cox survival model by likelihood ratio decomposition of model fit. **b-i**, Kaplan-Meier survival curves for top contributing blood factors.

Extended Figure 4: Vitals contribute to mortality risk

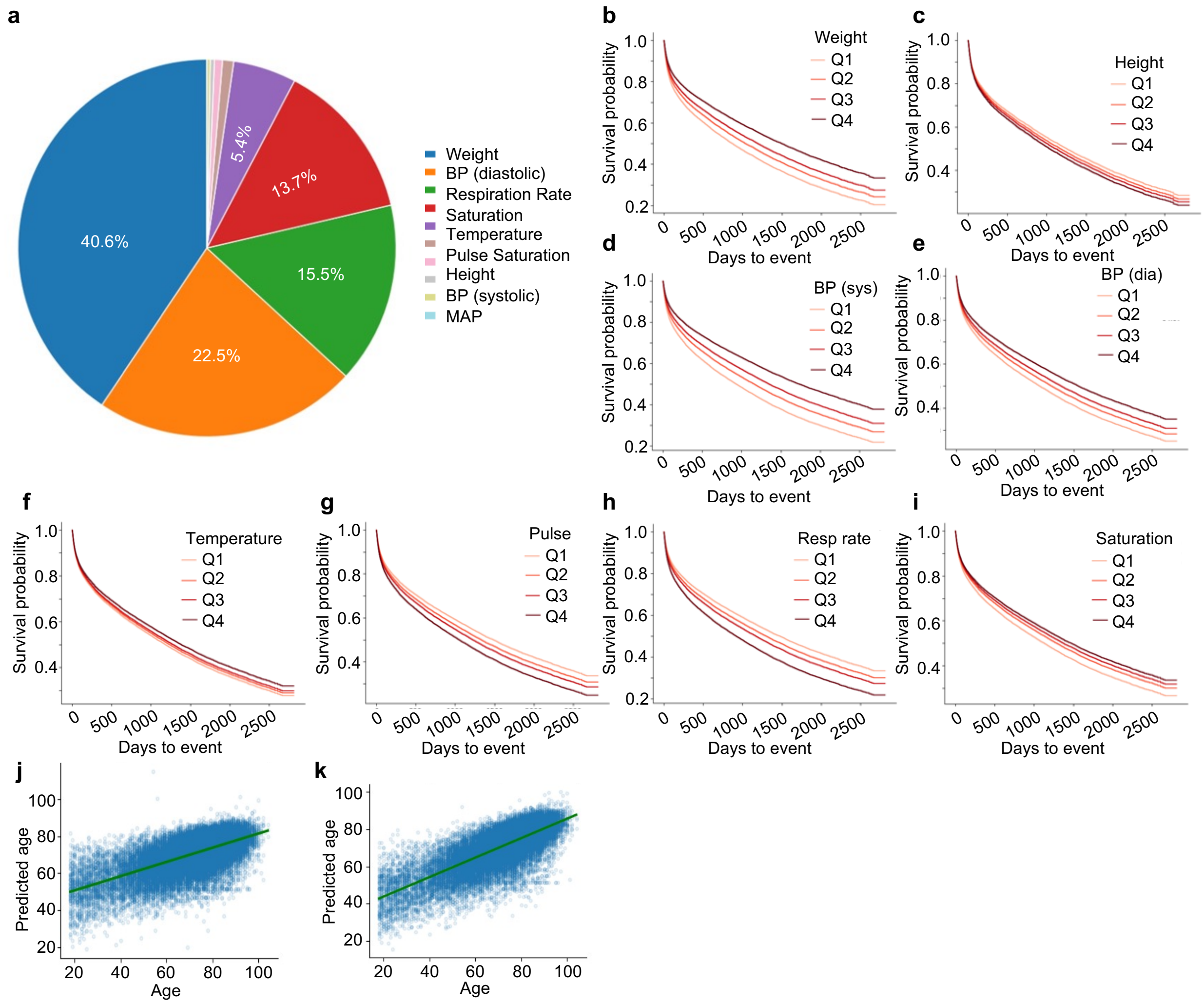


**a**, Breakdown of vital factors contributing to risk in a Cox survival model by likelihood ratio decomposition of model fit. **b-i**, Kaplan-Meier survival curves for top contributing vitals. **j**, Age predicted from blood factors (MAE=9.71, RMSE=12.75, n=27660). **k**, Age predicted from blood factors and vitals (MAE=8.26, RMSE=10.92, n=27660).

#### Extended Figure 5: Face regions and color contribute to mortality prediction

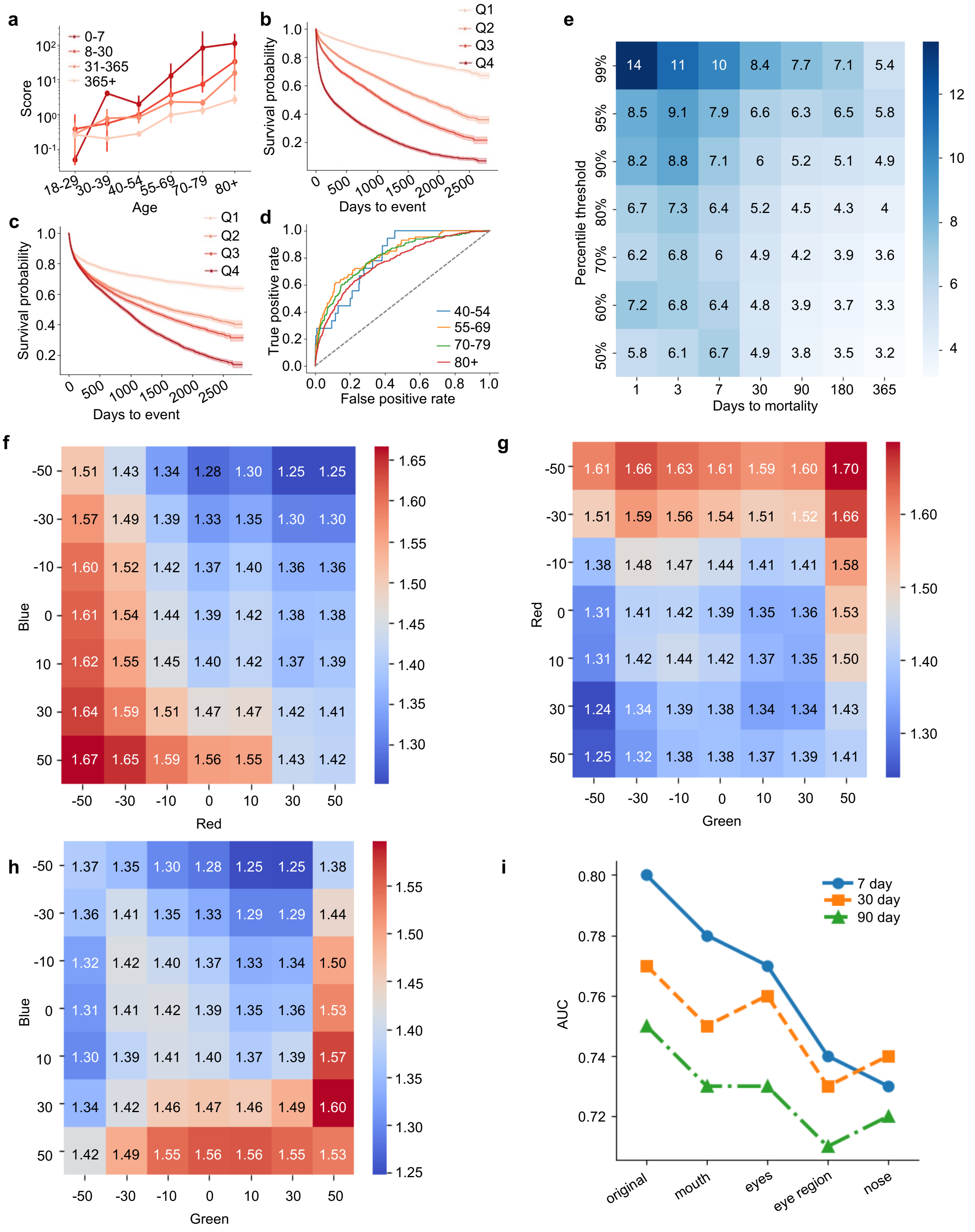


**a**, Mean risk score by age range, stratified by survival days. **b**, Kaplan-Meier survival curves for quartiles of risk scores. **c**, Kaplan-Meier survival curves by quartile of age-adjusted risk scores. **d,** ROC performance by age group for 7-day survival time (40-54: AUC=0.80, 55-69: AUC=0.83, 70-79: AUC=0.81, 80+: AUC=0.77). **e,** Odds ratios based on risk scores of upper quantiles vs lower for a set of time points. **f-h**, Heatmaps of risk scores from predicting images shifts in color channel intensities. **i**, AUC for several timepoints where facial features are occluded.
